# Yoga and Breathing Training in Patients with Heart Failure and Preserved Ejection Fraction: A Randomized Clinical Trial

**DOI:** 10.1101/2025.09.03.25335058

**Authors:** Carla Pinheiro Lopes, Luiz Claudio Danzmann, Daniel Umpierre, Ruy Silveira Moraes, Douglas Santos Soares, Santiago Alonso Tobar Leitão, Alexandre Silva Campos Filho, Flávio Depaoli, Luciano Pinto Guimarães, Gaspar Chiappa, Paulo José Cardoso Vieira, Jorge Pinto Ribeiro, Andreia Biolo

## Abstract

**BACKGROUND:** Patients with heart failure with preserved ejection fraction (HFpEF) usually have physical effort intolerance, and may present inspiratory muscle weakness (IMW). Yoga is known to have significant benefits on cardiovascular and respiratory health. However, the effects of yoga and breathing techniques on HFpEF have not been reported.

**METHODS AND RESULTS:** This study is a multicenter, randomized 1:1:1, outcome-blind, parallel-group, no-inferiority trial. Thirty-two patients with previous diagnosis of HFpEF (EF ≥ 50%) 45-75 years old, were randomized and allocated into groups: yoga group (Y = 11), breathing techniques (BT = 11) or the control group (C = 10) to evaluate the effects of an in-person program of 8-week of yoga and specific breathing techniques on inspiratory muscle responses, functional capacity, distinct features of the autonomic nervous system, natriuretic peptides, diastolic function, and quality of life. Data were analyzed using the Generalized Estimates equation model (GEE- GZLM). Yoga training resulted in 44.1% improvement in inspiratory muscle strength (Pth_max_) [19 (7-29) *vs* 2(-2 to 5) in control group, (p < 0.05). Breathing techniques increase heart rate variability (HRV) (827.6 ± 46.7ms vs 936 ± 51.4 ms, in control group,( p < 0.05). There was neither improvement in PI_max_, functional capacity nor in parameters of diastolic function.

**CONCLUSIONS:** In patients withHFpEF with and without IMW, yoga training was feasible and promoted increased respiratory muscle strength. Further studies should better address possible benefits on functional parameters. CLINICAL TRIAL REGISTRATION: URL: https://www.clinicaltrials.gov; NCT03028168; Protocol study: Trials. 2018;7:19:405. https://doi.org/10.1186/s13063-018-2802-5

## INTRODUCTION

Most patients with heart failure (HF) have limitations on physical activities due to the occurrence of dyspnea and peripheral muscle fatigue^1^. Many of them show respiratory muscle weakness (RMW) and/or deconditioning of skeletal muscles, which are involved in the metabolic dynamics of global movement, as well as an increased work of ventilatory muscles during hyperpnea^2^. In this context, additional factors, such as a decrease in resistance and maximal inspiratory pressure (PI_max_), are associated with decreased capacity response to exercise in patients with HF. Changes in these parameters are also associated with worsening of quality of life and disease progression^3,6^.

Inspiratory muscle training (IMT) appears to have satisfactory effects on functional class, inspiratory muscle strength and endurance, with increases in submaximal and maximal functional capacity and quality of life in HF patients with reduced ejection fraction (HFrEF)^6–96,7,9,10^. However, little attention has been given to ventilation, although evidence that breathing characteristics (frequency, amplitude and regularity) markedly affect beat-to-beat cardiovascular variability^11–13^. A recent study has compared the efficacy of various lifestyle modifications and ranked smoking cessation and adherence to yoga programs among the most effective changes in cardiovascular disease prevention^14^.

In physical work and specific respiratory techniques, the Yoga ventilatory control, characterized by complete diaphragmatic mobilization during the abdominal, thoracic and clavicular phases, may optimize respiratory volume and thus increase alveolar ventilation and perfusion. In addition, it has been reported that very slow and moderately slow yoga breathing creates significant changes in the frequency bands of heart rate variability (HRV) during and after exercise in healthy men and women^15,16^. Currently, yoga physical practices has been recommended in risk reduction programs^14^ and cardiovascular rehabilitation^17,18^.The primary outcome of this study was to assess the efficacy of yoga and breathing techniques on maximum inspiratory pressure (PI_max_) in patients with HFpEF. Therefore, this study was conducted to test the hypothesis that an 8-week yoga program and breathing techniques would lead to PI_max_ improvements in HFpEF when compared to the control group.

## METHODS

### Trial Design

The present study is a multicenter, randomized 1:1:1, outcome-blind, non-inferiority, parallel-group study, and was conducted at two specialized HF clinics (HF Clinic at Hospital de Clínicas de Porto Alegre (HCPA), RS, Brazil, and the HF ambulatory at Hospital ULBRA, Canoas, RS, Brazil), between August 2012 and March 2017. The study was registered in the Brazilian clinical trials platform - REBEC (RBR-64 mbnx) and in the international directory of clinical trials - Clinical Trials (NCT03028168). The protocol was published in *Trials*^19^.

### Participants

#### Inclusion and exclusion criteria

Adult patients aged from 45 to 75 years diagnosed with HFpEF, functional capacity class II and III, and who were being treated at a specialized HF clinic were eligible. HF diagnosis was established through medical history (signs and symptoms), echocardiographic findings (left ventricular ejection fraction ≥ 50%)^20,21^, and medical records confirming management for HF.

Exclusion criteria were unstable angina, myocardial infarction, or cardiovascular surgery within the previous 3 months, active orthopedic or infectious disease, and treatment with steroids, hormones, or cancer chemotherapy. Additionally, pulmonary disease (forced vital capacity < 80% predicted and/or forced expiratory volume for 1 s < 70% predicted)^5,22^, significant mitral or aortic valve diseases, record of exercise-induced asthma, and active smoking were also exclusion criteria. After selection, the discontinuous criteria were decompensated HFpEF, more than two consecutive absences in the intervention groups, and expressed willingness to discontinue at any time of the study or who did not sign the informed consent form.

### Interventions

Therapeutic interventions were given for two groups using different techniques for respiratory frequency and a control group without exercise therapy. While conducting the respiratory training, the interventors initially used a headset in one ear, with the sound of a metronome. Both interventions lasted 45 min per session and a standardized 7 min final relaxation was performed at the end of each intervention protocol.

All interveners were master’s, doctoral and undergraduate students from the health field. All collaborators were trained to perform their functions. The intervention videos are publicly available (https://osf.io/t7926/).

### Yoga (Y) – active breathing technique

Active protocol with yoga body movements (àsanas) performed along with respiratory technique without contentions, current and vigorous (ujjayi), observing a respiratory frequency of 15-20 respiratory cycles per minute. This session lasted around 45 min (Table S1).

### Yoga – passive breathing technique (pranàyama – BT)

Passive protocol, seated patient, no significant body movements. Yoga breathing technique with alternate nostril breathing (viloma pranàyama) uses diaphragmatic breathing, both current and combined with inspiratory and expiratory retention, observing a slow respiratory frequency of between 5 and 8 respiratory cycles per minute. This session lasted approximately 45 min (Table S2).

### Control group (C)

Patients were oriented to keep their pharmacological routine and daily activities. They were scheduled to return to the hospital for post-testing after 8 weeks of randomization. After final assessment, all patients, including those in the control group, were invited to participate in the study breathing activities at the outpatient wards of this trial.

### Outcomes

The primary endpoint was inspiratory muscle strength assessed by the maximal inspiratory pressure (PI_max_). Secondary endpoints included (1) vagal activity in resting and exercising (HRV); (2) QoL Minnesota scores as a specific inventory for patients with HF; (3) volumetric ratios of left atrial and diastolic pressure gradients on echocardiography; and (4) changes in BNP/NT-Pro-BNP tests between pre- and post-intervention measurements.

### Test Protocols Pulmonary function tests

#### Respiratory muscle strength

Respiratory muscle strength was determined using the Pi_max_ and PE_max_ measurements and the MVD-300 manovacuometer (Microhard System, Globalmed, Porto Alegre, Brazil). During the procedure, the patients remained seated at rest, upright, their nostrils occluded with a nasal clip and their mouths properly set in the mouthpiece, avoiding air leaks. To measure PE_max_, the individual was asked to inhale at the level of total lung capacity (TLC), followed by a maximal expiratory effort. PI_max_ was obtained by performing an inspiration, using TLC, generating maximal inspiratory effort. The patient performed three to six attempts on the test, and the evaluator was instructed to record all the values obtained for PI_max_ and PE_max_ (cmH_2_O). The PI_max_ and PE_max_ values were recorded using the highest value obtained by the individual and should not surpass the second highest value by 10%. During the assessments, the evaluator strongly encouraged the individual to give his or her maximum using incentive phrases.

When PI_max_ value was < 70% of the predicted value for gender and age, it was considered as a decrease in inspiratory muscle strength or inspiratory muscle weakness (IMW)^22^.

### Progressive load test of inspiratory muscles

The inspiratory muscle strength was determined using the modified progressive loading protocol of Martyn et al^23^. Inspiration was initiated with a loading of 50% of PI_max_, and then the load was increased by 10% of the PI_max_ every three minutes. The patient ventilated continuously until the inspiratory valve of the equipment could not be opened.

The resistance index was determined by the maximal sustained inspiratory pressure (Pth_max_) for at least 60 s in the last stage, and will be expressed as a percentage of the maximal inspiratory pressure (Pth_max_/PI_max_,%)^24^.

At each stage, heart rate (HR), Respiratory rate (RR), oxygen saturation by pulse oximetry (Sp) (Datex-Ohmeda, 3800 Oximeter, Louisville, USA) and the dyspnea index by the perception scale of BORG were monitored.

### Constant load test

To test the inspiratory muscle strength, the patient ventilated against 80% of Pth_max_ for the maximum tolerated time. The improvement in the individual’s performance was evaluated by the total duration of the test in seconds (s)^25^.

### Heart rate variability analysis

Heart rate variability indices were calculated from twenty-four-hour ECG recordings obtained with a SEER Light digital recorder (GE Medical Systems Information Technologies, Milwaukee, WI). The recorded data were analyzed using a MARS 8000 analyzer (Marquete Medical Systems, Milwaukee, WI) as previously described^26^. In short, time domain indices were calculated according to recommendations of the European Society of Cardiology and North American Society of Pacing and Electrophysiology^27^. The following 24 h indices were calculated: mean of all normal RR intervals, SD of all normal RR intervals, root mean square of successive differences of adjacent RR intervals, and percentage of successive differences between normal adjacent RR intervals above 50 ms (RR, pNN50, RR, rMSSD, SDNN). Only patients in sinus rhythm were included.

### Quality of Life Questionnaire

Quality of life (QOL) was assessed using the Minnesota Living With Heart Failure questionnaire^28^. Overall scores as well as the separate effects of physical and psychological perceptions on QOL were analyzed.

### Six-Minute Walk Test (TC-6)

The submaximal functional capacity was recorded by means of the TC-6, in which the greatest distance that the individual walked within a fixed time interval of six minutes was measured^29^. Subjects were instructed to perform the test according to the attached protocol (NYHA).

Blood pressure (BP) and Respiratory rate (RR) were measured at the beginning and end of the test. The HR, peripheral oxygen saturation (measured by digital oximeter) and the BORG dyspnea scale were recorded at the beginning of the test, every minute and at the end of the test. The predicted distance for the individual was estimated using Enright and Sherrill’s equations^30^.

### Echocardiogram

The usual and additional echocardiographic measurements, such as volumetric gradients and diastolic parameters, were analyzed in the selection of eligible patients (LVEF calculated using the Teichholz method – EF ≥ 50%)^21^ and using two-dimensional echocardiograms after the intervention (PHILLIPS IEE 33).

### BNP and Pro-BNP

Due to adherence to the method of modified analysis at the hospital of origin, two different methodologies were used to measure BNP. Initially, blood samples of the natriuretic peptide type B (BNP), clinical sample by serum, with chemiluminescence analysis method (ADVIA CENTAUR SIEMENS).

The N-terminal precursor test of B-type natriuretic peptide (NT-proBNP) was adopted using the sandwich-type electrochemiluminescence analysis (COBAS E601- ROCHE). In the pre and post tests, the same biomarker (BNP or NT-proBNP) was employed^31,32^. Classifications were analyzed for the diagnosis of HFPEF and for comparison after the interventional treatment. For the analysis of the two methodologies, the variables were transformed into a Z-scale in order to set same metric values. For each method, the mean and standard deviation were calculated by the formula Z = xi - <u>𝑥</u> / si , where "i" is the subject, "x" is the BNP value observed, "<u>𝑥</u>" is the BNP mean of the method, and “s” is the standard deviation of each method.

### Sample Size

Based on previous studies, the sample size was calculated using inspiratory muscle pressure as endpoint^33^. Considering a difference among treatments (effect size) of 15 cm H_2_O and a standard deviation (SD) of 12 cm H_2_O in the PI_max_, representing a 1.2 ratio (effect size/SD), using an α = 0.05 and power of 80%, 9 patients would have to be included per group. In addition, considering a potential loss in patient follow-up of between 10% and 20%, the sample was set to 11 patients per group.

### Randomization

Patients who met the eligibility criteria were invited to participate in the study; those who accepted to participate signed a written informed consent form. Following the conclusion of pre-intervention tests, the researcher informed the biostatistics center at HCPA, which is in charge of the randomization list, and participants were allocated to either yoga (active) or yoga (passive) or control group in a 1:1:1 ratio using a pre-generated simple randomization list. The allocation sequence was blinded from researchers and outcome assessors by a central.

### Blinding

The researchers were divided according to their specific role in this study as (1) interveners – those performing the protocol interventions (blinded for outcomes, but not for groups); (2) medical appraiser – responsible for performing clinical tests (blinded for groups, but not for outcomes); and (3) analysts – those responsible for the statistical analyses (blinded for both groups and outcomes). The individuals were instructed to avoid talking with the research team about protocol intervention and clinical trials.

### Deviation from Registered Protocol

The authors acknowledge the following deviations from our registered protocol: (a) omission of VO2 max results through ergospirometry that were not considered safe for reproduction due to failure detected in the equipment cell, at the end of the collection phase (b) reports on the secondary result measurement (Pth) linked to the PI_max_ result, as it is included in the main objective of this manuscript, due to increased interest in these end points.

### Statistical Methods

A descriptive analysis was performed and data were expressed as absolute and relative frequency, besides mean and standard deviation or quartiles, accordingly. The treatment groups were compared using the generalized estimating equation, specific for repeated measurements, in order to compare the effects (means) across the three groups and the two times, in addition to the group × time interaction. The generalized estimating equation matrix of robust estimator covariance and exchangeable work correlation matrix were used if normal distribution is found and will be analyzed by an identity binding function. In contrast, if an asymmetrical distribution is found, data were analyzed using a gamma distribution linked to a logarithmic function. When significant, the factors under study were compared by Bonferroni’s post-hoc test. Correlations will be described by Pearson’s or Spearman’s test. PASW18 (version 18.0, SPSS, Chicago, Illinois, USA) will be used in this analysis.

## RESULTS

### Patients

Between August 2012 and March 2017, 841 patients with HF were screened for the study. After verification of the inclusion and exclusion criteria, 42 patients were recruited; however, because of additional exclusions, only 36 were randomized. Among the 12 patients randomized to the “Yoga group” - Y, 1 did not complete the training protocol because it was not possible to attend sessions. Among the 13 patients allocated to “Breathing Techniques group” - BT, 1 was excluded because of excessive absences in protocol, and 1 developed HFrEF criteria. Among the 11 patients allocated to “Control Group” - C, one did not complete pre-protocol tests. Patients were encouraged to adhere through telephone contacts. Therefore, 32 patients (Table 1) completed the protocols and tests (Figure 1). The initial protocol was kept unchanged during the follow-up of the study. No interim analysis was performed.

**Figure 1.**
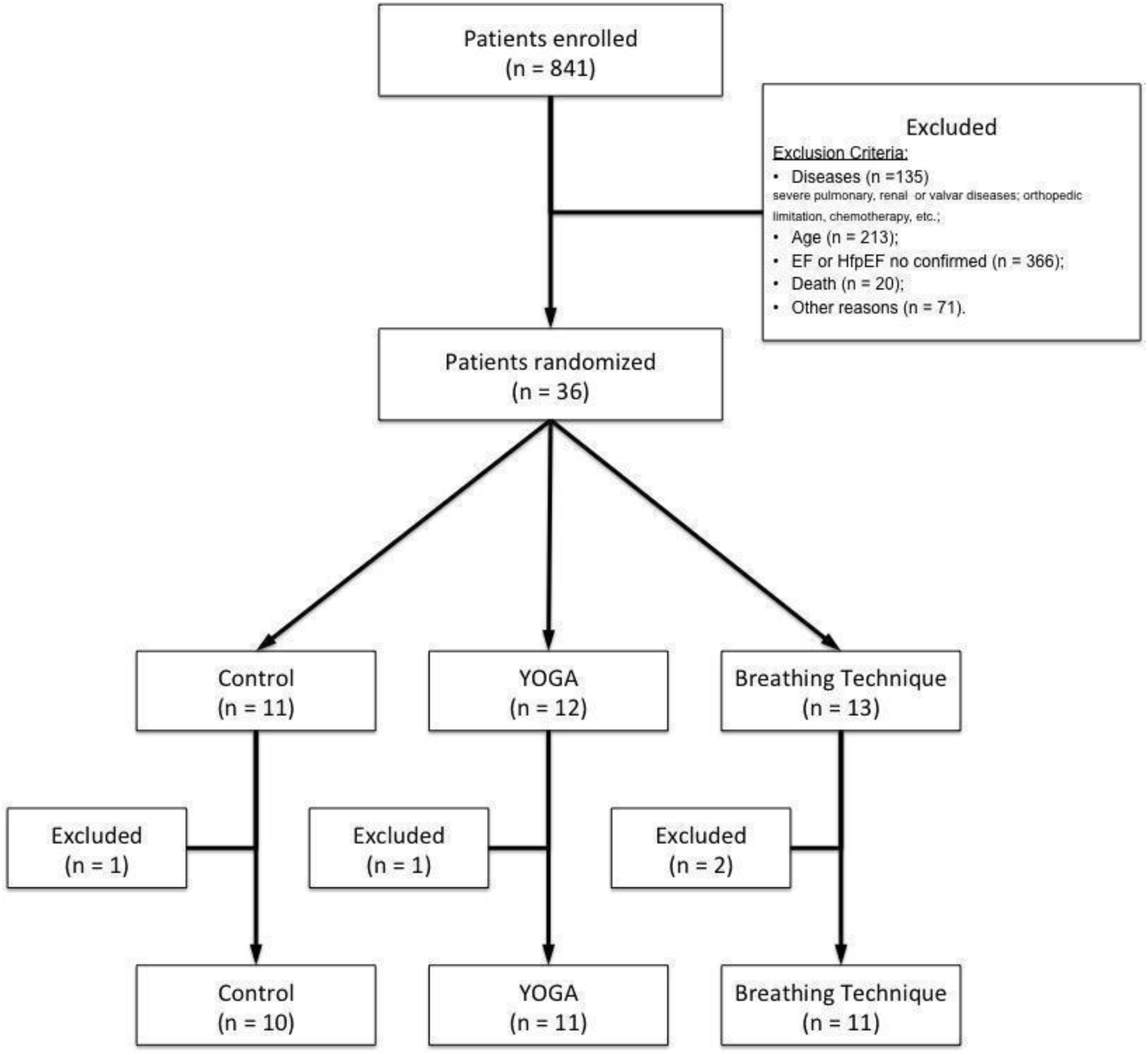
Flow diagram of study. Number of participants who were initially screened, randomized and included in the final analysis. EF, ejection fraction; HFpEF, heart failure with preserved ejection fraction.

**Table 1.** Baseline characteristics of patients randomized to yoga, breathing techniques, and control groups. SD,Standard deviation; BMI, Body mass index; HF, Heart failure; AF, Atrial fibrillation; IMW, Inspiratory muscle weakness; ACEi, Angiotensin-converting enzyme inhibitors.

| Characteristics | Yoga (n =11) | Respiratory (n = 11) | Control (n = 10) |
| --- | --- | --- | --- |
| Female (male) | 10 (1) | 6 (5) | 6 (4) |
| Age (years;SD) | 66.7 ± 6 | 64.9 ± 8 | 62.0 ± 6 |
| BMI (kg/m<sup>2</sup>;SD) | 33.9 ± 5 | 32.7 ± 8 | 33.8 ± 5 |
| Obesity (n;%) | 7 (63.6) | 5 (45.5) | 8 (80) |
| Sedentary (n;%) | 9 (81.8) | 7 (63.6) | 6 (60) |
| Functional class (NYHA) |  |  |  |
| I | 0 | 0 | 0 |
| II | 6 | 7 | 10 |
| III | 5 | 4 | 0 |
| HF etiology (n) |  |  |  |
| Ischemic | 5 | 2 | 2 |
| Non-ischemic (n) | 6 | 9 | 8 |
| AF (n;%) | 6 (54.5) | 0 | 4 (40) |
| IMW (n) | 2 | 5 | 4 |
| Mitral Failure (mild/moderate)(n;%) | 3 (27.2) | 2 (18.2) | 2 (20) |
| Hypertension (n;%) | 11 (100) | 9 (81.8) | 9 (90) |
| Diabetes (n;%) | 3 (27.3) | 5 (45.4) | 3 (30) |
| Dyslipidemia (n;%) | 9 (81.8) | 8 (72.7) | 7 (70) |
| Ejection fraction (%;SD) | 68.7 ± 1.7 | 67.0 ± 1.5 | 66.7 ± 2.1 |
| Drugs (n;%) |  |  |  |
| Beta-blockers | 8 (72.7) | 8 (72.7) | 7 (70) |
| ACEi | 8 (72.7) | 5 (45.5) | 8 (80) |
| Digitalic | 4 (36.3) | 1 (9.1) | 0 |
| Diuretics | 10 (90.9) | 6 (54.5) | 7 (70) |
| Antiplatelets | 7 (63.6) | 5 (45.4) | 6 (60) |

### Inspiratory muscle function tests

After 16 weeks, yoga and respiratory exercises induced improvement in PI_max_ (20 cm H_2_O for Y group, 19 cm H_2_O for BT group, and 6 cm H_2_O for C group) with a significant PI_max_ for comparison between the time before and after the intervention within the BT group. This difference occurred within the BT group but not when compared between the Y and C groups. (Figure 2). Although sustained inspiratory muscle strength significantly increased (p < 0.05) in the Y group at the end of the incremental load test (Pth_max_), Pth_max_/PI_max_ (%) function increased when compared to the BT group (19 cm H_2_O; 14 cm H_2_O). This increase in ventilatory quality in the BT group, although it may be clinically relevant, was not statistically significant (Table 2 and Figure 2 ).

**Figure 2.**
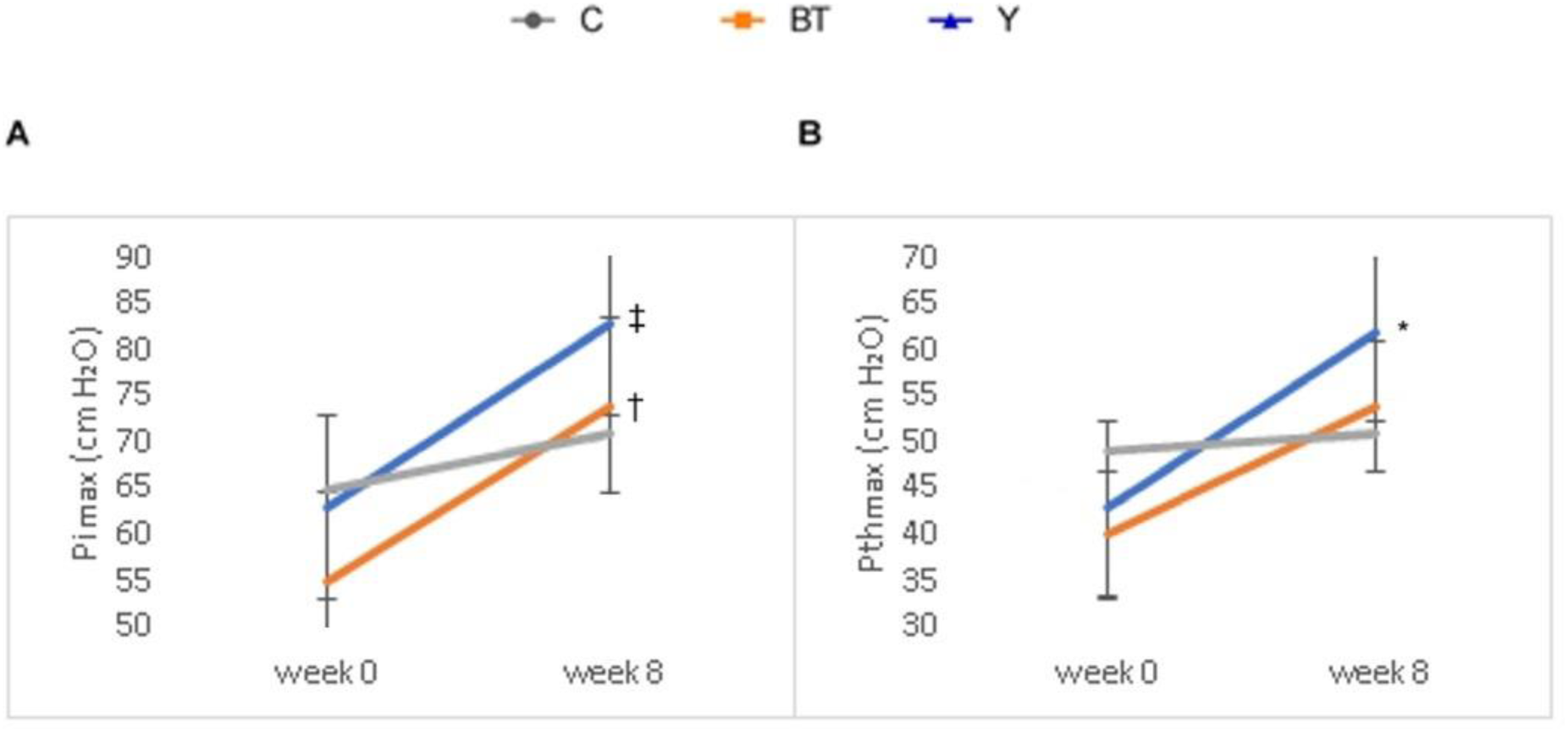
Main outcome at baseline and follow-up. (A). PImax measured using a manovacuometer (B). Pthmax measured using a powerbreathe K5. Expressed as mean ± SD. * p < 0.05 for group-time interaction; † p < 0.001 for time comparison; ‡ p < 0.05 for time comparison.

**Table 2.**
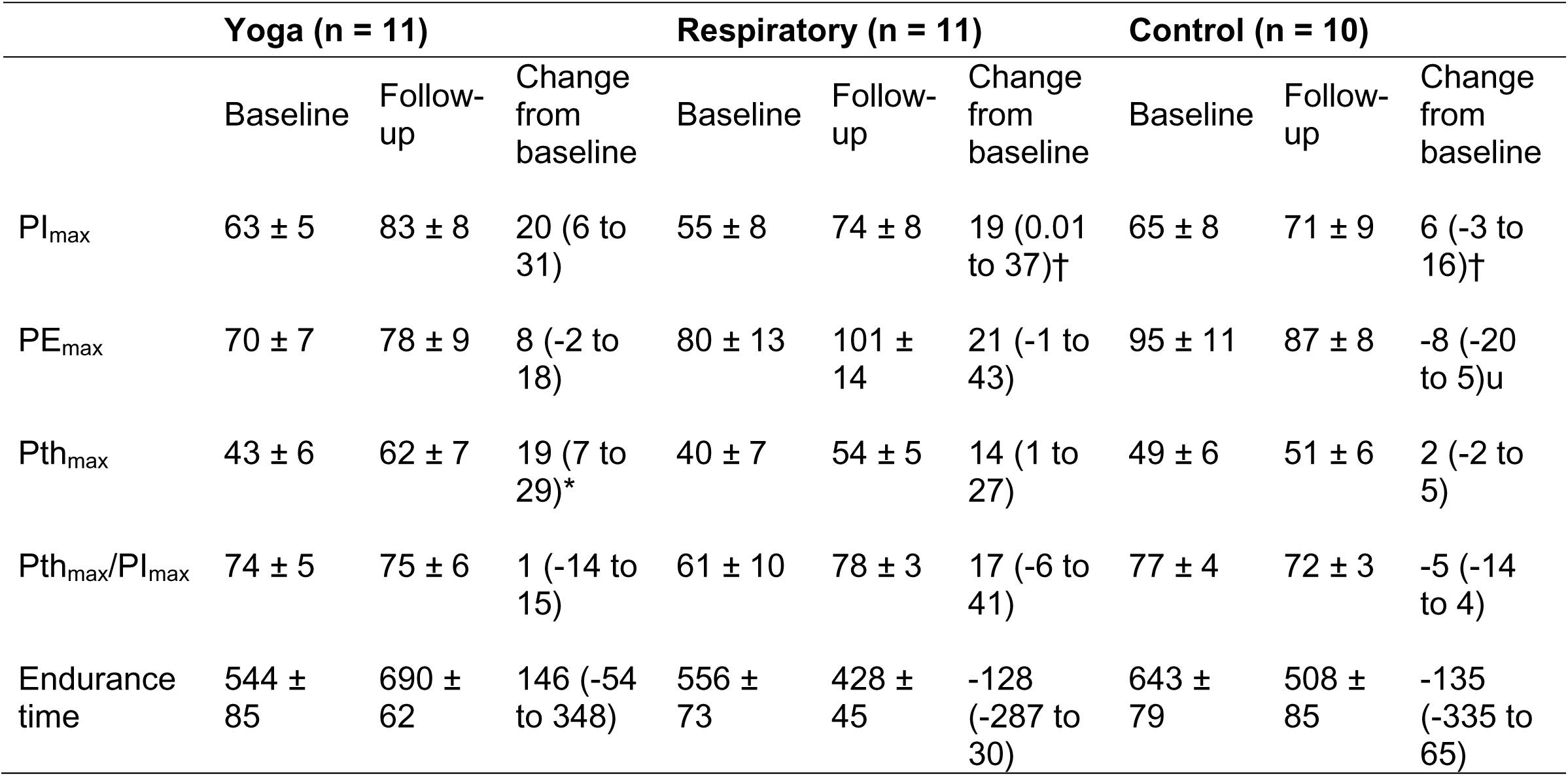
Inspiratory muscle function testing data at baseline and post training compared among yoga, respiratory techniques and control groups. Data expressed as mean ± SD. PI_max_, maximum inspiratory pressure; PE_max_, maximum expiratory pressure; Pth_max_, maximum inspiratory pressure and time sustained for incremental test. * p < 0.05 for group-time interaction; † p < 0.001 for time comparison; ‡ p < 0.05 for time comparison.

The endurance time sustained in the inspiratory muscle presented a statistical trend (p = 0.07) and the yoga group reached a higher mean when compared to the respiratory techniques group in post-training (p <0.05), (Table 2). As to PI_max_, a statistical difference was observed in the time comparison post-intervention period in the BT (p < 0.001), where mean values were higher too in the Y group , but not in the control group (0.196) (Figure 2).

### Autonomic responses

The time domain parameters (Table 3) of HRV, including mean RR interval (p = 0.000), NN50 count divided by the total number of all NN intervals (pNN50) (p = 0.02), and square root of the mean of the sum of the squares of differences between adjacent NN intervals (rMSSD) (p = 0.04) increased in the Breathing Techniques group, when compared to the other groups or to the time of intervention (Table 4). In the frequency domain analysis, power of low-frequency (LF), power of high frequency (HF) as well as in the time domain, the standard deviation variable of all NN intervals (SDNN) did not attain statistical significance (Figure 3).

**Figure 3.**
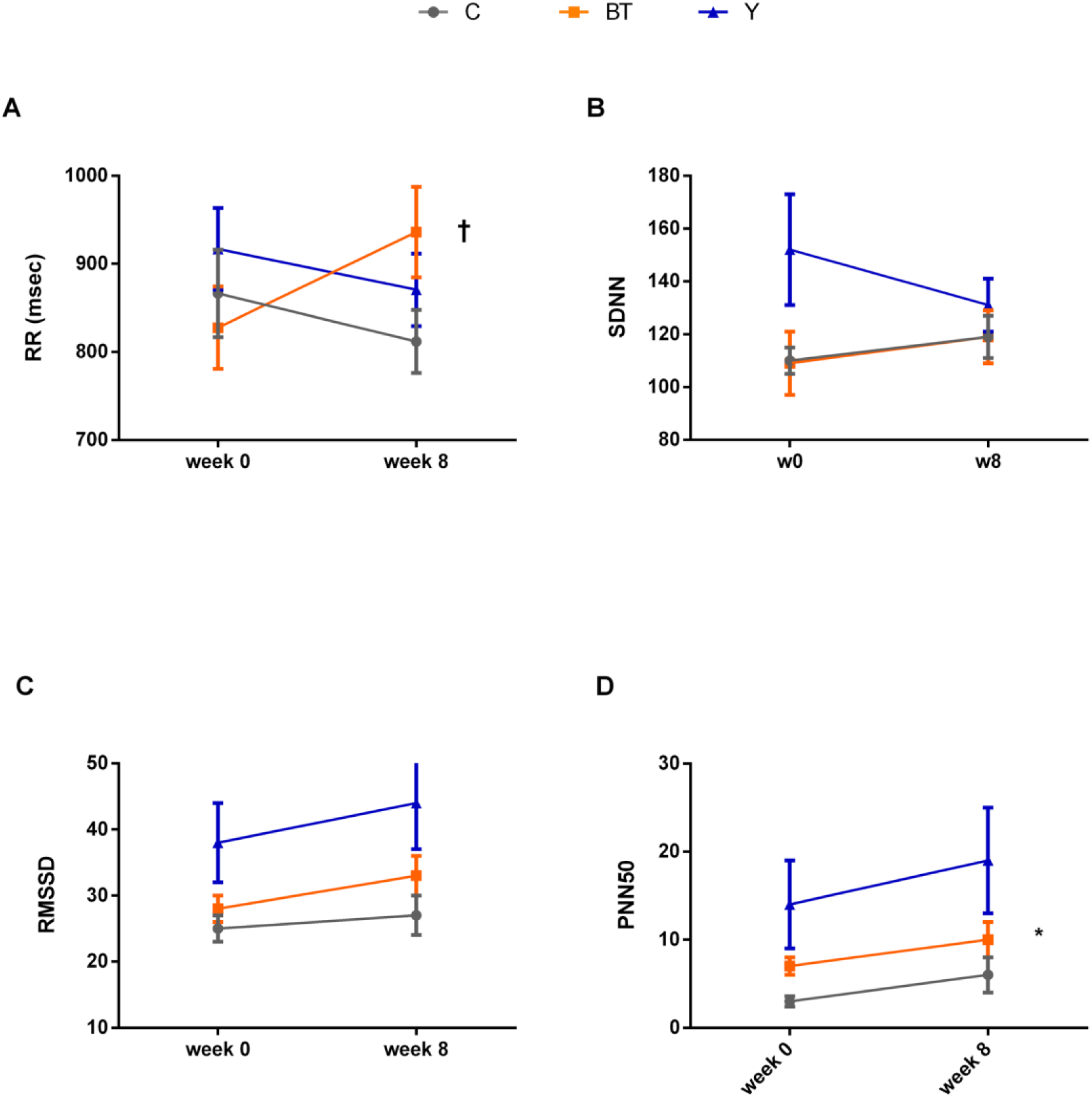
HRV parameters (RR,SDNN,rMSSD,pNN50, illustrations A, B, C, D, respectively) outcomes at baseline and follow-up. Values are expressed as mean ± SD. Δ, difference delta from follow-up and baseline. * p < 0.05 for different means in comparisons; † p < 0.001 for group-time interaction.

**Table 3.**
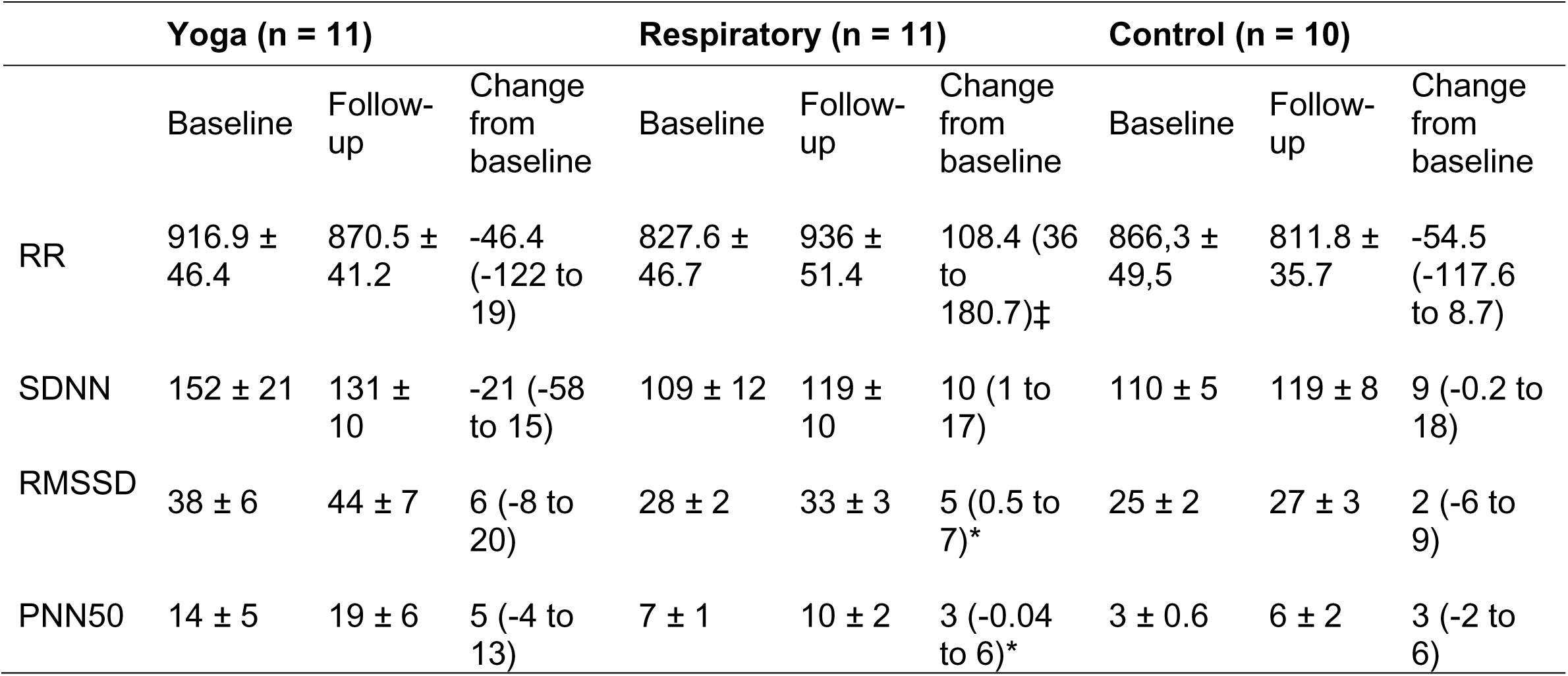
Distribution of time domain means throughout the 24h in yoga group, breathing techniques group, and control group. RR: interval variability; pNN50 (%): SD of all normal RR intervals; rMSSD: square root of the mean of the sum of the squares of differences between adjacent NN intervals; SDNN: standard deviation of all NN intervals. Values are expressed as mean ± SD. *p<0.05 for different means in comparisons; ‡ p< 0.05 for group-time interaction

**Table 4.** Other secondary outcomes at baseline and follow-up. Values are expressed as mean ± SD. HR, heart rate; Borg, subjective effort scale; BNP, brain natriuretic peptide; ∅ LVEDD, left ventricle end diastolic diameter; ∅ LVESD, left ventricle end systolic diameter, EF, ejection fraction; LVM, left ventricle mass; E/e’, early diastolic peak velocity and diastolic peak velocities of the mitral annulus ratio. * p < 0.05 for different means in comparisons; † p < 0.001 for time comparison; ‡ p < 0.05 for time comparison.

|  | Yoga (n = 11) |  |  | Respiratory (n = 11) |  |  | Control (n = 10) |  |  |
| --- | --- | --- | --- | --- | --- | --- | --- | --- | --- |
|  | Baseline | Follow-up | Change from baseline | Baseline | Follow-up | Change from baseline | Baseline | Follow-up | Change from baseline |
| 6- min. walk (m) | 387 ± 12 | 432 ± 22 | 45 (8 to 82)*‡ | 442 ± 31 | 471 ± 22 | 29 (-8 to 66)‡ | 453 ± 33 | 497 ± 21 | 44 (-32 to 121)‡ |
| Expected distance | 421 ± 10 | 425 ± 11 | 4 (-2 to 10) | 454 ± 16 | 452 ± 16 | -2 (-8 to 5) | 457 ± 16 | 492 ± 37 | 35 (-35 to 105) |
| HR (bpm) (baseline) | 64 ± 3 | 62 ± 3 | -2 (-8 to 3) | 70 ± 3 | 67 ± 3 | -3 (-7 to 2) | 69 ± 3 | 70 ± 5 | 1 (-6 to 9) |
| HR (bpm) (end) | 92 ± 9 | 84 ± 3 | -9 (-27 to 10) | 99 ± 7 | 87 ± 6 | -12 (-28 to 4) | 93 ± 9 | 101 ± 8 | 8 (-8 to 24) |
| Borg | 6 ± 05 | 5 ± 0.4 | -1 (-2 to -0.06)‡ | 7 ± 0,4 | 7 ± 0.5 | -0,5 (-1 to 0.8)‡ | 7 ± 0.4 | 7 ± 0.5 | -0.3 (-2 to 1)‡ |
| BNP (ng/L) | 0.5 ± 0.4 | 0.8 ± 0.4 | 0.3 (-0.24 to 0.9) | -0.4 ± 0.2 | -0.3 ± 0.2 | 0,1 (-0.13 to 0.36) | -0.4 ± 0.2 | 0.3 (-0.2 to 0.8) | 0.3 (-0.2 to 0.8) |
| Aortic diameter (mm) | 3.1 ± 0.1 | 31 ± 0.07 | 0.05 (-0.1 to 0.2) | 3.3 ± 0.1 | 3.2 ± 0.1 | -0.1 (-0.1 to 0.05) | 3.4 ± 0.1 | 3.5 ± 0.1 | 0.1 (-0.004 to 0.13) |
| Left atrium diameter (mm) | 4.3 ± 0.1 | 4.2 ± 0.1 | -0.1 (-0.2 to 0.1) | 4.2 ± 0.1 | 4.3 ± 0.1 | 0.1 (-0.02 to 0.2) | 4.1 ± 0.03 | 4.1 ± 0.07 | 0.02 (-0.1 to 0.1) |
| Ø LVEDD (mm) | 4.9 ± 0.1 | 4.7 ± 0.1 | -0.2 (-0.3 to 0.05) | 4.7 ± 0.1 | 4.4 ± 0.1 | -0.3 (-0.5 to 0.02) | 4.9 ± 0.1 | 5 ± 0.1 | 0.1 (-0.2 to 0.3) |
| Ø LVESD (mm) | 3 ± 0.1 | 2.8 ± 0.1 | -0.2 (-0.004 to 0.3) | 29 ± 0.1 | 2.8 ± 0.1 | -0.1 (-0.2 to 0.04) | 3.1 ± 0.1 | 3.2 ± 0.1 | 0.1 (-0.2 to 0.3) |
| EF (%) | 68.7 ± 1.7 | 70.2 ± 2 | 1.5 (2.1 to -5.1) | 67 ± 1.5 | 65.7 ± 1.3 | -1.3 (-3.3 to 0.8) | 66.7 ± 2.1 | 65.7 ± 2.8 | -1 (-6.6 to 4.5) |
| LVM (g) | 202 ± 15 | 189 ± 13 | -13 (-28 to 3) | 213 ± 26 | 216 ± 31 | 3 (-16 to 21) | 228 ± 8 | 217 ± 16 | -11 (-42 to 21) |
| Mass/body surface (g/m<sup>2</sup>) | 111 ± 6 | 107 ± 7 | -4 (-12 to 2) | 111 ± 10 | 108 ± 13 | -3 (-11 to 6) | 114 ± 7 | 121 ± 9 | 7 (-3 to 17) |
| E/e' | 12 ± 1.4 | 12 ± 1.5 | 0.05 (-0.09 to 1.1) | 10 ± 0.9 | 10 ± 0.8 | 0.1 (-0.7 to 0.9) | 10 ± 1.1 | 11 ± 1.5 | 1 (-0.8 to 2.3) |

### Other outcomes

On the 6-minute walk test, all patients, regardless of the group, showed shorter distances at baseline for gender, age, and body weight. After training, the mean meters covered increased in groups Y, BT, and C, with respective Δ, 45 m, 28 m and 44 m (Table 5).

**Table 5.** Quality of life scores. Δ, difference delta from follow-up and baseline. MLHF, minnesota living with heart failure scores

|  | Yoga (n = 11) |  |  | Respiratory (n = 11) |  |  | Control (n = 10) |  |  |
| --- | --- | --- | --- | --- | --- | --- | --- | --- | --- |
|  | Baseline | Follow-up | Change from baseline | Baseline | Follow-up | Change from baseline | Baseline | Follow-up | Change from baseline |
| Emotional | 15 ± 7 | 9 ± 2 | -6 (-9 to -3) | 7 ± 6 | 6 ± 1 | -1 (-3 to 1) | 1- ± 5 | 7 ± 2 | -3 (-6 to 0) |
| Physical | 19 ± 2.9 | 11 ± 2.2 | -8 (2.2 to 14.1) | 16 ± 3.3 | 12 ± 2.7 | -4 (0.8 to 37.4) | 18 ± 3.6 | 15 ± 3.2 | -3 (1.9 to 16.3) |
| MLHF | 43.5 ± 5.2 | 25.9 ± 3.5 | -17.6 (-30 to -4) | 38 ± 5.6 | 27.7 ± 4.8 | -10.3 (-18.7 to -1.8) | 36.4 ± 5.5 | 32.2 ± 2.9 | -4.2 (-15.3 to 6.9) |

The outcomes related to Borg, BNP and echocardiography showed no differences across the groups. MLHF score results decreased in the 3 groups; however, in group Y, Δ showed higher reduction (-17.6), followed by BT group (-10.3) and C group (-4.2). As to physical and emotional domains, group Y showed a higher score decrease (respectively - 8; - 6); group BT (- 4; - 1) and group C (- 3; - 1) showed similar reduction between time before and after intra-group intervention (Table 5).

### Subgroup analysis

#### IMW

The Respiratory variables were adjusted by IMW (n = 11). After adjustment, data have not changed significantly, confirming IMW independence to the Respiratory outcomes obtained.

### Diabetes in autonomic analyses

The adjustment for diabetes (n = 11) in the autonomic model showed no difference when compared to previous analyses. In addition, in SDNN analysis, there was a group*time interaction trend (p = 0.06) that was not shown before remission in diabetes.

## DISCUSSION

In this randomized clinical trial conducted in patients with HFpEF, yoga and breathing techniques programs for an eight-week training period improved inspiratory muscle strength and endurance as well as heart rate variability. These improvements occurred regardless of factors that decrease ventilatory or autonomic efficiency, such as weak inspiratory muscles or diabetes. However, there were no changes in functional capacity or diastolic function parameters between the intervention and control groups.

In accordance with previous studies, PI_max_ increased in groups Y and BT. In group Y, the Pth_max_, which indicates the Sustained Muscle Inspiratory Pressure (SMIP) or inspiratory muscle inspiratory, showed significant gains during the incremental load test. However, in the constant load resistance test, no significant differences were observed between groups. It is worth noting that these tests rely on different primary metabolic pathways (glycolytic vs. oxidative, respectively) which may have been influenced by the interval and glycolytic demands from yoga protocol. These findings underscore improvements in cardiorespiratory variables following YG intervention in patients with HFpEF.

According to previous studies in patients with HFrEF^7,9,10,34–36^, IMT improves inspiratory muscle strength and endurance. Meanwhile, our findings revealed that patients with HFpEF exhibited significant increases in PI_max_ and inspiratory pressure (Pth outcomes), even without a targeted inspiratory training regimen. This result should be underscored because the present study was based on nasal breathing whereas other studies used mouth breathing for inspiratory training, which are closely compatible with the specificity of the assessment tests.

Inspiratory muscle function has traditionally been evaluated using PI_max_. However, PI_max_ is rarely required in daily physical activities, which typically depend on repeated respiratory muscle contractions demanding both strength and endurance. The SMIP offers a more comprehensive assessment of this endurance capacity, reflecting the ability of respiratory muscles to maintain strength over time. Prior investigations have demonstrated that SMIP is strongly correlated with clinical outcomes in COPD and provides superior discriminatory power compared to PI_max_ indices for predicting ventilatory work^37,38^. These findings highlight the importance of assessing inspiratory muscle endurance in future studies, particularly in populations like HFpEF, where adaptations may not be solely attributed to maximum force production.

The clinical aspects of HFpEF highlight exercise intolerance as particularly severe compared to other cardiovascular diseases^39^, often accompanied by varied degrees of pulmonary and systemic congestion^40^, functional limitations, autonomic vagal dysfunction, obesity, and marked chronotropic incompetence^41^. These multifactorial limitations significantly impact the ventilatory process. This finding aligns with the understanding that HFpEF often involves diminished ventilatory capacity, whether or not chronic obstructive pulmonary disease (COPD) is present. Poor recruitment of accessory muscles to support diaphragm function likely contributes to respiratory inefficiency in these patients, particularly during prolonged or non-interval exercise.

In the assessment of HRV, our findings indicate that slower respiratory rates (RR) performed with alternate nasal blocks and respiratory restrictions during BT increase HRV in patients with HFpEF. Similarly, beta-blockers and ACEI/ARBs reduce neurohormonal activation, morbidity, and mortality in HF^42,43^. In other health conditions, previous studies have shown that right nostril yoga breathing increases blood pressure measures, while left nostril breathing decreases them, suggesting yoga may influence blood pressure in healthy individuals^44^. In patients with hypertension, slower breathing techniques over three months reduced blood pressure^45^, and combining yoga and breathing improved symptoms, arrhythmia burden, mental health, and quality of life in patients with atrial fibrillation^46^. These findings corroborate with our results of improved autonomic function, as indicated by HRV indexes, and reinforces the applicability of breathing techniques across distinct clinical conditions.

Some limitations of the study need to be considered. The study’s short intervention period and limited session frequency may have restricted functional capacity improvements. Larger sample sizes could enhance sample representativeness and reproducibility for outcomes such as the 6-minute walk test, Borg scale, and QoL scores^28^.

Further studies on yoga and breathing might focus on biochemical changes, central vagal stimulation, and changes in HR induced by respiratory rate, and these might clarify parameters of exercise intolerance and contribute to the reduction of morbidity and mortality in HFpEF.

Finally, in patients with HFpEF yogic training with physical postures, carried out with breathing techniques and without additional loads, promoted an increase in SMIP. Training with the yoga-pranayama breathing technique, with ventilation conducted more slowly, showed an increase in vagal activity, which may derive clinical benefits for patients with HFpEF. Notably, we did not observe improvement in the functional capacity of these patients, which may warrant further research on how such findings may be mechanistically integrated.

## Data Availability

Datasets will be shared upon request.

https://osf.io/t7926/

## Data sharing

Datasets will be shared upon request.

## Non-standard Abbreviations and Acronyms

AF: Atrial Fibrillation
BP: Blood Pressure
BT: Breathing Technique
Group C: Control Group
COPD: Chronic Obstructive Pulmonary Disease
CVD: Cardiovascular disease
DBP: Diastolic Blood Pressure
DM: Diabetes Mellitus
ETT: Exercise Tolerance
Test FC: Functional Capacity
HFrEF: Heart Failure with Reduced Ejection Fraction
HRV: Heart Rate Variability
IMT: Inspiratory Muscle Training
IMW: Inspiratory Muscle Weakness
MBP: Mean Blood Pressure
PI_max_ = MIP: Maximal static Inspiratory Pressure
Pth_max_ = SMIP: Maximal Inspiratory Pressure sustained for 1 min during incremental test
QoL: Quality of Life
RCT: Randomized Clinical Trial
RF: Respiratory Frequency
RMW: Respiratory Muscle Weakness
RR: Respiratory Rate
RT: Respiratory Training
SAH: Systemic Arterial Hypertension
SBP: Systolic Blood Pressure
Y: Yoga Group

## Supplemental Material

**Table S1.**
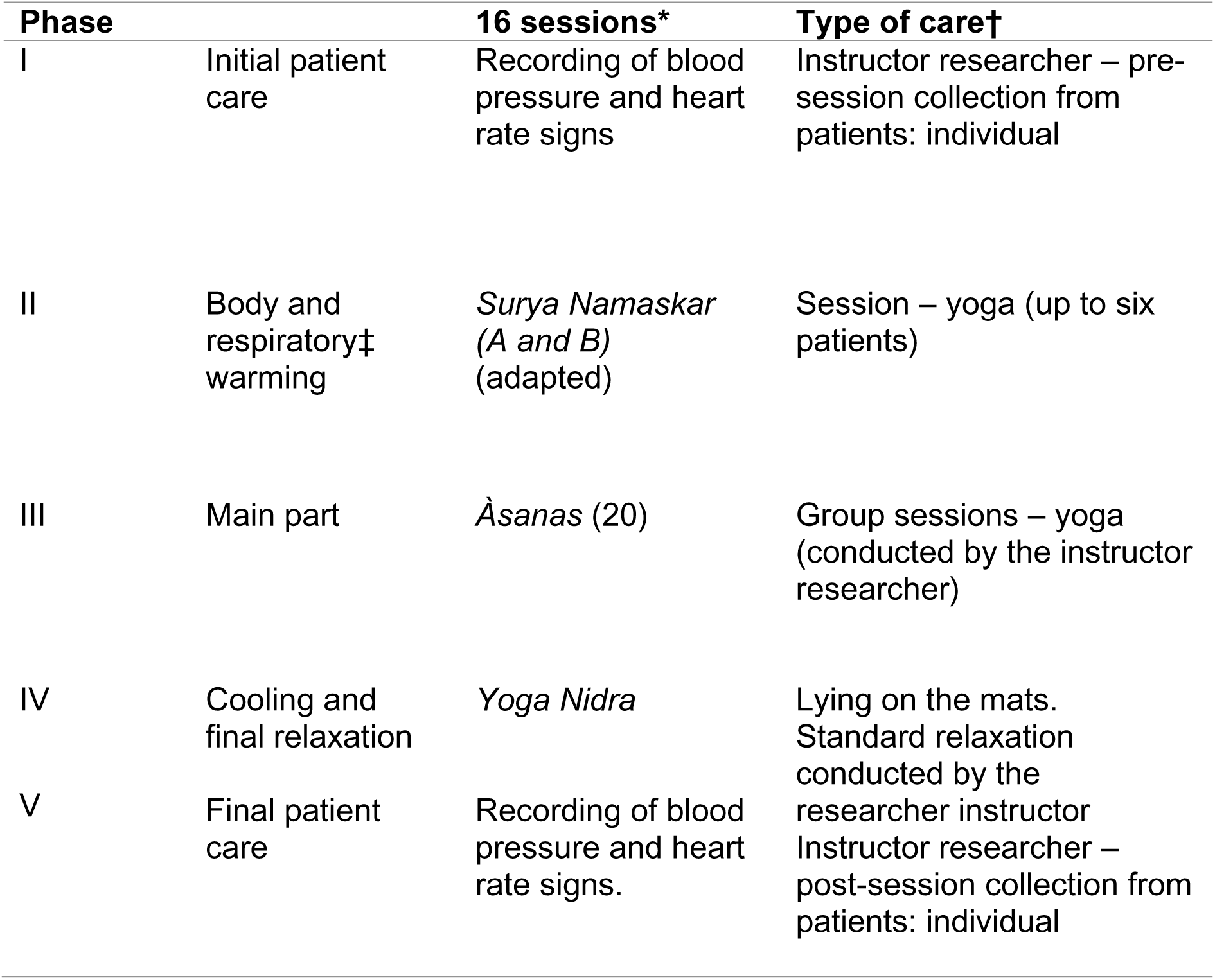
Yoga group: five phases of the session in intervention programme. * Sessions of approximately 45 minutes at 1 hour, twice a week; † Physiatrics room (20 m^2^, approximately): 7 mats; 6 chairs; 1 sphygmomanometer; 1 stethoscope; 1 support stretcher. ‡ Breath: *Ujjayi pranàyama*, frequency 15-20 breaths per minute (bpm).

| Phase |  | 16 sessions* | Type of care† |
| --- | --- | --- | --- |
| I | Initial patient care | Recording of blood pressure and heart rate signs | Instructor researcher – pre-session collection from patients: individual |
| II | Body and respiratory‡ warming | *Surya Namaskar (A and B)* (adapted) | Session – yoga (up to six patients) |
| III | Main part | *Āsanas* (20) | Group sessions – yoga (conducted by the instructor researcher) |
| IV | Cooling and final relaxation | *Yoga Nidra* | Lying on the mats. Standard relaxation conducted by the researcher instructor |
| V | Final patient care | Recording of blood pressure and heart rate signs. | Instructor researcher – post-session collection from patients: individual |

**Table S2:** Breathing technique group: five phases of the session in intervention programme. * Sessions of approximately 45 minutes at 1 hour, twice a week; †Physiatrics room (20 m^2^, approximately): 7 mats; six chairs; 1 sphygmomanometer; 1 stethoscope; 1 support stretcher. ‡ 7 sets of *anulom vilom/nadi shodhana pranayama* (8 breaths each); frequency around 5-8 breaths per minute (bpm).

| Phase |  | 16 sessions* | Type of care† |
| --- | --- | --- | --- |
| I | Initial patient care | Recording of blood pressure and heart rate signs | Instructor researcher – pre-session collection from patients: individual |
| II | Warming up: Breathing conscientization | Complete breathing exercises (Diaphragmatic, middle and chest) - Leid | Lying on the mats. Diaphragmatic and complete breathing conducted by the researcher instructor |
| III | Warming up of the upper body muscles - chair | Diaphragmatic release exercises | Session – intervention with seated patients (up to six patients) |
| IV | Main part | *Anulom vilom/nadi shodhana pranayama* technique‡ | Alternate nostril breathing, without and with *kumbhaka* – inhalation and/or exhalation retentions (conducted by the instructor researcher) |
| V | Cooling and final relaxation | *Yoga Nidra* | Lying on the mats. Standard relaxation conducted by the researcher instructor |
| VI | Final patient care | Recording of blood pressure and heart rate signs. | Instructor researcher – post-session collection from patients: individual |

